# Integrating Machine Learning for Propensity Score Matching and Causal Inference: A Causal Forest Approach to Assessing the Impact of Maternal Education on Antenatal Care Utilization

**DOI:** 10.1101/2025.07.10.25331269

**Authors:** Sultan Mahmud, Md Mohsin, Razia Mustani, Mt. Sonia Akter

## Abstract

**Background:** While maternal education is linked to antenatal care (ANC) use, its causal effect remains uncertain. This study applies a machine learning approach, Causal Forests, to estimate the causal impact of maternal education on adequate ANC utilization in Bangladesh.

**Methods:** We analyzed data from 6,815 ever-married women aged 15–49 years who had a live birth within five years preceding the 2022 Bangladesh Demographic and Health Survey (BDHS). The outcome was adequate ANC utilization, defined as receiving four or more ANC visits from skilled providers. Maternal education was dichotomized as <secondary vs. ≥secondary education. To estimate causal effects, we employed a machine learning–based propensity score matching approach using K-nearest neighbors (KNN), followed by treatment effect estimation using Causal Forests. We also assessed treatment heterogeneity across subgroups and conducted a Rosenbaum bounds sensitivity analysis to evaluate robustness to unmeasured confounding.

**Results:** While a strong crude association was observed between maternal education and ANC use (83.4% vs. 16.6%, *p* < 0.001), the estimated Average Treatment Effect (ATE) after matching was modest and statistically non-significant (ATE = 0.006, 95% CI: –0.002 to 0.016, *p* = 0.17). However, significant heterogeneity in Individual Treatment Effects (ITEs) was detected across subgroups, with higher effects among women aged 20–40 at first birth, urban residents, those with media exposure, and those with wealthier or more educated spouses. The sensitivity analysis indicated moderate robustness to hidden bias.

**Conclusions:** Maternal education alone may not significantly impact ANC use once socioeconomic and contextual factors are accounted for. However, its benefits are amplified among specific subgroups, suggesting the need for integrated and context-sensitive maternal health interventions. Advanced machine learning methods can enhance causal inference and inform equity-oriented policy strategies.

## Introduction

Maternal mortality remains a critical global public health issue, with an estimated 810 women dying each day due to complications related to pregnancy and childbirth (1). The vast majority of these deaths, about 99%, occur in low- and middle-income countries (LMICs), with Southern Asia and Sub-Saharan Africa bearing the highest burden, accounting for approximately 66% and 20% of maternal deaths, respectively (2). Most of these fatalities are preventable through timely access to essential, low-cost maternal health interventions during pregnancy, childbirth, and the postpartum period (2).

Antenatal care (ANC) is a cornerstone of maternal and newborn health, providing essential preventive, diagnostic, and therapeutic services that significantly improve pregnancy outcomes (3). Achieving health-related Sustainable Development Goals (SDGs) has led the Government of Bangladesh to prioritize universal access to antenatal care (ANC) services. As a result, the proportion of women receiving ANC at least once during pregnancy rose significantly from 34% in 2000 to 82% by 2017/18 (2). However, the uptake of the recommended four or more ANC visits remains relatively low. According to the 2022 Bangladesh Demographic and Health Survey, only 39.8% of women received four or more ANC visits, with wide variations by wealth, education, and region (4).

Maternal education has emerged as one of the most influential determinants of maternal health service uptake, including antenatal care (ANC) (2,5). Literature shows that educated women are more likely to identify pregnancy concerns early, seek prompt care from trained experts, and successfully navigate complicated healthcare systems (1,6–8). Despite these observed associations, establishing causal links between maternal education and ANC utilization remains challenging due to confounding factors (e.g., socioeconomic status, geographic location) inherent in observational data (9).

When confounding is present, traditional regression techniques frequently fail to estimate causal effects, especially in cross-sectional data. By mimicking a quasi-experimental design, propensity score approaches provide a solution that enables the estimation of treatment effects while controlling for observed covariates (1,10). This study addresses this gap by integrating advanced machine learning methods for propensity score estimation with Causal Forest modeling to estimate the causal impact of maternal education on adequate ANC utilization in Bangladesh.

Traditional causal inference methods like Propensity Score Matching (PSM) have been employed to address confounding by constructing comparable treatment (educated) and control (non-educated) groups (10–13). However, PSM faces limitations, including sensitivity to model misspecification, difficulties in high-dimensional settings, and an inability to model heterogeneous treatment effects (11,14).

Recent advances integrate machine learning with causal inference to overcome these constraints. The Causal Forest method, an extension of random forests, estimates Conditional Average Treatment Effects (CATE) by:

1. Using nuisance models to adjust for confounders via local centering (14).
2. Employing “honest” causal trees that partition data to maximize treatment effect heterogeneity (14).
3. Aggregating predictions across trees in an ensemble to robustly estimate individualized treatment effects (14).

This approach efficiently handles non-linear relationships and high-dimensional covariates while quantifying effect heterogeneity—capabilities beyond conventional PSM (14).

This study leverages the Causal Forest framework to rigorously assess the causal impact of maternal education on ANC utilization, using data from the nationally representative 2022 Bangladesh Demographic and Health Survey (BDHS). BY integrating machine learning methods with causal inference, we aim to uncover nuanced patterns of treatment heterogeneity that are often overlooked in conventional analyses. The findings are expected to contribute methodologically to the advancement of causal inference in health services research and, practically, by informing targeted public health interventions aimed at reducing maternal health inequities in Bangladesh and similar low-resource settings.

## Methods

### Data source

This study utilized publicly available data from the Bangladesh Demographic and Health Survey (BDHS)-2022 (15), a nationally representative cross-sectional survey conducted by the National Institute of Population Research and Training (NIPORT) with technical assistance from ICF under the DHS Program. The BDHS collects comprehensive demographic, socioeconomic, and health-related information through structured interviews with women of reproductive age (15–49 years) across all administrative divisions of Bangladesh using a two-stage stratified cluster sampling design.

The sampling frame was based on the Integrated Multi-Purpose Sampling Master Sample developed by the Bangladesh Bureau of Statistics (BBS) using the 2011 population census. It included detailed information on enumeration areas (EAs), such as geographic location, type of residence (city corporation, other urban, or rural), and the estimated number of residential households. Each EA was accompanied by a sketch map outlining its geographic boundaries.

Administratively, Bangladesh is divided into eight divisions—Barishal, Chattogram, Dhaka, Khulna, Mymensingh, Rajshahi, Rangpur, and Sylhet—which are further subdivided into zilas, upazilas, and smaller administrative units. These divisions formed the basis for stratification into urban and rural areas during sampling.

In the first stage of sampling, 675 EAs were selected with probability proportional to size, including 237 from urban areas and 438 from rural areas. A complete household listing was then conducted within each selected EA to create a sampling frame for the second stage. In the second stage, a systematic sample of 45 households per EA was selected. Among these, 30 households were randomly chosen for the long-form questionnaire, which was administered to all eligible ever-married women aged 15–49, while the remaining 15 households received a shorter version of the questionnaire.

Within the long-form households, 15 were selected for biomarker collection, which included height and weight measurements for children under age five and ever-married women aged 15–49. From these, 8 households were further selected for extended anthropometric and blood pressure measurements among ever-married women aged 50 and above, never-married women aged 18 and above, and all men aged 18 and older. This rigorous sampling design ensured reliable and representative estimates of key demographic and health indicators across urban and rural areas and all eight administrative divisions of Bangladesh.

### Study participants

The analytical sample comprised ever-married women aged 15–49 years who had at least one live birth in the five years preceding the survey. To ensure consistency in the analysis, only the most recent live birth was considered for women with multiple births during this period.

### Exposure (treatment) variable

The primary exposure variable in this analysis was maternal education, operationalized as a binary indicator. Women were classified according to their highest level of formal education: those with less than secondary education (no education, incomplete primary, or completed primary) were coded as 0, while those who had completed secondary school or higher were coded as 1.

### Outcome variable

The outcome variable was adequate antenatal care (ANC) utilization, defined in accordance with World Health Organization recommendations. Specifically, adequate ANC was considered as having received four or more ANC visits from skilled providers during the most recent pregnancy, coded as a binary variable (anc_4plus), where 1 indicated receipt of four or more visits and 0 indicated fewer than four.

### Covariates (potential confounders)

A range of covariates was included to control for potential confounding in the relationship between maternal education and ANC utilization. These were selected based on prior literature and subject-matter expertise (1,16,17). Age at first birth (age_first_birth) was categorized as ≤19 years (coded as 1) or 20–40 years (coded as 2). Religion (mother_religion) was classified as Islam (1) or other religions (2, including Hinduism, Buddhism, Christianity, etc.). Place of residence (mother_residence) distinguished urban (1) from rural (2) settings.

The number of living children (num_living_child) was grouped as 0 (no children), 1 (1–2 children), or 2 (3–10 children). Administrative division (division) reflected the respondent’s geographic region, covering all eight administrative divisions of Bangladesh. Wealth index (wealth_index) provided a categorical measure of household socioeconomic status, ranging from poorest to richest, and is a standard control in DHS-based analyses (18).

Husband’s education (hudb_education) categorized as less than secondary (1) or secondary and higher (2). Media exposure (media_exposure) was a binary variable indicating whether the woman reported exposure to at least one form of media—newspaper, radio, or television. Short birth interval (short_birth_interval) was defined as an interval of less than 24 months between births and recoded as 1 (<24 months) or 2 (≥24 months).

Finally, perceived distance to a health facility (distance_problem) was captured as a binary variable, indicating whether the respondent considered distance a significant barrier (1 = big problem, 0 = not a big problem).

### Propensity score estimation

To reduce confounding and approximate a quasi-experimental design, propensity scores (PS) were estimated to represent the probability of receiving the treatment—defined here as having secondary or higher maternal education—conditional on observed covariates. Formally, the propensity score P𝑆(𝑉) is defined as:

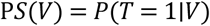

where 𝑇 = 1 indicates treatment (secondary or higher education), and V is the vector of observed covariates. Estimating P𝑆(𝑉) allows balancing of covariates between treated and control groups, thereby reducing confounding bias in observational studies (19,20).

In addition to logistic regression, we compared multiple machine learning algorithms to identify the best-performing model that could enhance model flexibility and potentially improve the accuracy and robustness of propensity score estimation. The following models were employed to estimate propensity scores:

### Logistic Regression

Logistic regression is a probabilistic machine learning approach commonly used for classification tasks, including the estimation of propensity scores in causal inference studies. In this context, it models the probability of receiving the treatment (e.g., having secondary or higher education) as a function of observed covariates. By fitting a logistic function to the covariates, the model estimates the propensity score, defined as the conditional probability of treatment assignment given the covariates. The traditional formulation models the log-odds of treatment assignment as a linear combination of these covariates, allowing researchers to adjust for confounding and create balanced groups for comparison:

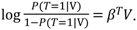

Where, 𝑃(𝑇 = 1|𝑉) represents the probability of being in the treatment group (i.e., having secondary or higher education), given the observed covariates 𝑉 and 𝛽 coefficient of 𝑉.

### Support Vector Machines (SVM)

Support Vector Machines (SVMs) are supervised learning algorithms that perform well in high-dimensional settings and can capture complex, non-linear decision boundaries. In the context of propensity score estimation, SVMs aim to classify individuals into treated and untreated groups by finding an optimal hyperplane that separates the two classes in a high-dimensional feature space. This hyperplane is chosen to maximize the margin between the groups, thereby improving generalizability. The resulting decision function reflects the boundary that best distinguishes treatment assignment based on observed covariates:

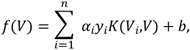

where 𝐾 is a kernel function, 𝑦_𝑖_ are class labels, and 𝛼_𝑖_ are learned coefficients, 𝑏 is the bias term.

### Random Forest

Random Forests are ensemble learning methods that aggregate the predictions of multiple decision trees, each trained on a different bootstrap sample of the data and a random subset of covariates. This approach helps reduce variance and improve predictive accuracy. In the context of propensity score estimation, each tree provides a predicted probability of treatment assignment based on the covariates, and the final propensity score estimate is obtained by averaging these probabilities across all trees. Mathematically, the estimated propensity score is given by:

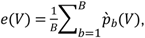

where 𝐵 is the total number of trees in the forest, and 𝑝_𝑏_(𝑉) is the predicted probability of treatment for individual 𝑉 from the where 𝑏^𝑡ℎ^tree. This averaging process results in a more stable and robust estimate of the propensity score compared to a single decision tree.

### Neural Networks

Neural networks, particularly feedforward multilayer perceptrons (MLPs), are capable of modeling complex, nonlinear relationships between covariates and treatment assignment. These models consist of multiple layers of interconnected nodes, each applying an activation function to the weighted sum of inputs. In propensity score estimation, the output layer typically uses a sigmoid activation function to produce probabilities:

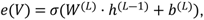

where, 𝜎 is the sigmoid function, 𝑊^(𝐿)^and 𝑏^(𝐿)^ are weights and biases at the output layer, and ℎ^(𝐿―1)^ are activations from the previous layer. Neural networks can approximate highly nonlinear and complex functions, making them flexible tools for estimating propensity scores in settings with intricate relationships among covariates (21).

#### K-Nearest Neighbors (KNN)

K-Nearest Neighbors is a non-parametric method that estimates propensity scores by averaging the treatment status of the 𝑘 most similar individuals (i.e., nearest neighbors) in the covariate space. The estimated propensity score is given by:

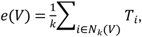

Where, 𝑁_𝑘_(𝑉) is the set of 𝑘 nearest neighbors of 𝑉. While simple and interpretable, the performance of KNN depends heavily on the choice of 𝑘 and the distance metric used.

#### Extreme Gradient Boosting (XGBoost)

XGBoost is a scalable and efficient implementation of gradient boosting, designed to handle complex prediction tasks. For propensity score estimation, XGBoost sequentially builds an ensemble of decision trees, where each new tree is fitted to the residuals (errors) of the previous model to refine the predictions. The output is the predicted probability of treatment given covariates. The model minimizes a regularized objective function of the form:

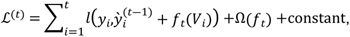

where, 𝑙 is a differentiable loss function, 𝑓_𝑡_ is the prediction from the tree added at iteration 𝑡, 𝑦 is the accumulated prediction up to iteration 𝑡 ― 1 and Ω(𝑓_𝑡_) is a regularization term that penalizes model complexity.

### Causal Forest model

Causal Forest is a machine learning method designed to estimate heterogeneous treatment effects at the individual or subgroup level, combining ideas from random forests and causal inference (22). It extends the traditional random forest by focusing not on predicting outcomes directly, but on estimating the Conditional Average Treatment Effect (CATE), denoted as 𝜏(𝑣), which varies with covariates 𝑣.

Causal Forest builds on Robinson’s transformation, which reformulates the treatment effect estimation as a residual-on-residual regression problem. For each observation 𝑖, the outcome 𝑌_𝑖_ and treatment indicator 𝑊_𝑖_ are centered by their respective nuisance functions: the outcome regression 𝑚(𝑉_𝑖_) and the propensity score 𝑆𝑃(𝑉_𝑖_). The transformed problem is to estimate 𝜏(𝑣) by minimizing the squared residuals:

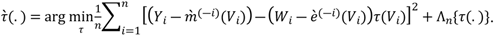

Here, 𝑚^(―𝑖)^ and 𝑒^(―𝑖)^are out-of-bag estimates, excluding observation 𝑖, and Λ_𝑛_is a regularization term controlling model complexity (22). This approach creates “pseudo-outcomes” (residuals) on which the forest is trained, allowing it to adaptively partition the covariate space to identify subgroups with differing treatment effects

### Model selection for propensity score estimation

To assess model performance, we focused primarily on the ability of each approach to achieve covariate balance between treatment groups after propensity score matching. Covariate balance was quantified using standardized mean differences (SMDs), with values below 0.1 considered indicative of acceptable balance (10). In addition to SMDs, we examined the overlap in propensity score distributions between the treatment and control groups using density plots, which visually demonstrate the extent to which the groups are comparable (23).

Further, we incorporated global balance measures such as distance-based metrics (for example, the Lévy distance) to capture distributional differences between matched groups (24). The efficiency of each matching procedure was also evaluated by recording the number of matched pairs retained after matching. Finally, the predictive accuracy of each model was assessed using the area under the receiver operating characteristic curve (AUC), which reflects the model’s ability to discriminate between treated and untreated subjects.

The optimal model was selected based on a composite assessment of these metrics. Specifically, the model that achieved the lowest standardized mean differences across covariates, the highest density overlap, the minimal adjusted distance imbalance, the largest number of matched pairs, and the highest AUC value was chosen for subsequent matching and treatment effect estimation.

### Statistical analysis

The sample characteristics were presented using frequency and percentage distributions. Statistical tests were conducted to examine the associations between the outcome and the treatment variable, as well as between the outcome and other covariates, in the unmatched data. Matching and treatment effect estimation were adjusted using the same set of covariates. Based on the estimated propensity scores, we performed 1:1 nearest neighbor matching with a caliper of 0.1 to create balanced treatment and control groups. To estimate heterogeneous treatment effects and the average treatment effect (ATE), we applied the Causal Forest algorithm. Specifically, we estimated the Average Treatment Effect (ATE), Conditional Average Treatment Effect (CATE), and Individual Treatment Effects (ITEs).

To assess whether the estimated individual treatment effects (ITEs) varied across population subgroups, we conducted a heterogeneity analysis using the ITEs derived from the causal forest model. For binary and categorical variables with two levels, we used Welch’s two-sample t-tests to compare mean ITEs between groups. For variables with more than two categories, we used one-way analysis of variance (ANOVA) to test for overall differences in mean ITEs across groups. To quantify the magnitude of subgroup differences, we reported Cohen’s d effect sizes for binary comparisons and eta squared (η²) for ANOVA models. To assess the robustness of the estimated treatment effects to potential hidden bias from unobserved confounding, we conducted a sensitivity analysis using Rosenbaum bounds. All analyses were conducted using R version 4.5.0 (2025-04-11 ucrt), and statistical significance was assessed at the 0.05 level.

## Results

### Sample characteristics

A total of 6,815 women were in unmatched (full) data (Table 1). Of them, 41.8% received adequate antenatal care (ANC ≥ 4 visits). Adequate ANC utilization was significantly higher among women with at least secondary education (83.4%) compared to those with lower education (16.6%) (p < 0.001). Similar patterns were observed for husbands’ education, with 69.9% of adequately cared-for women having spouses with higher education (p < 0.001).

**Table 1.**
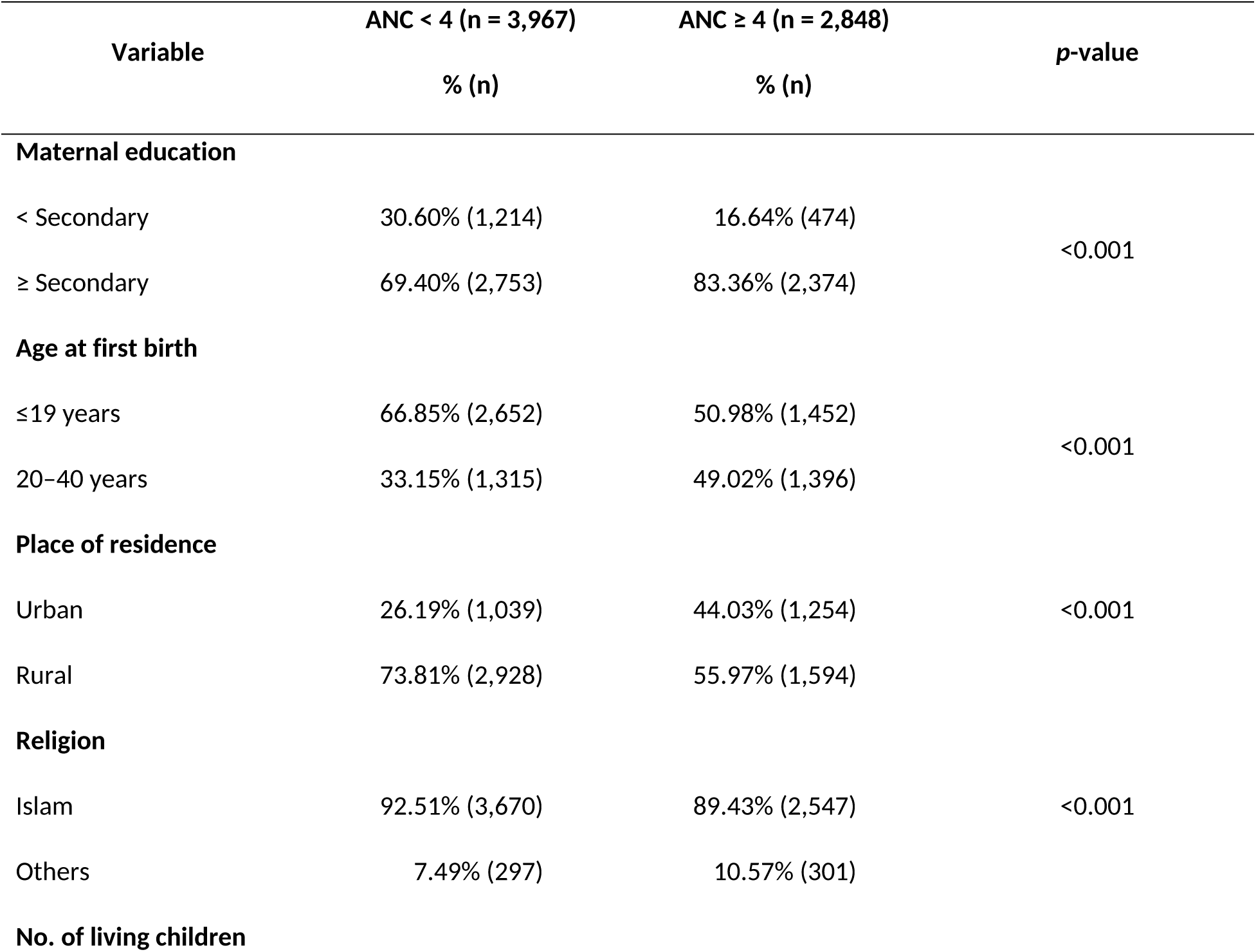
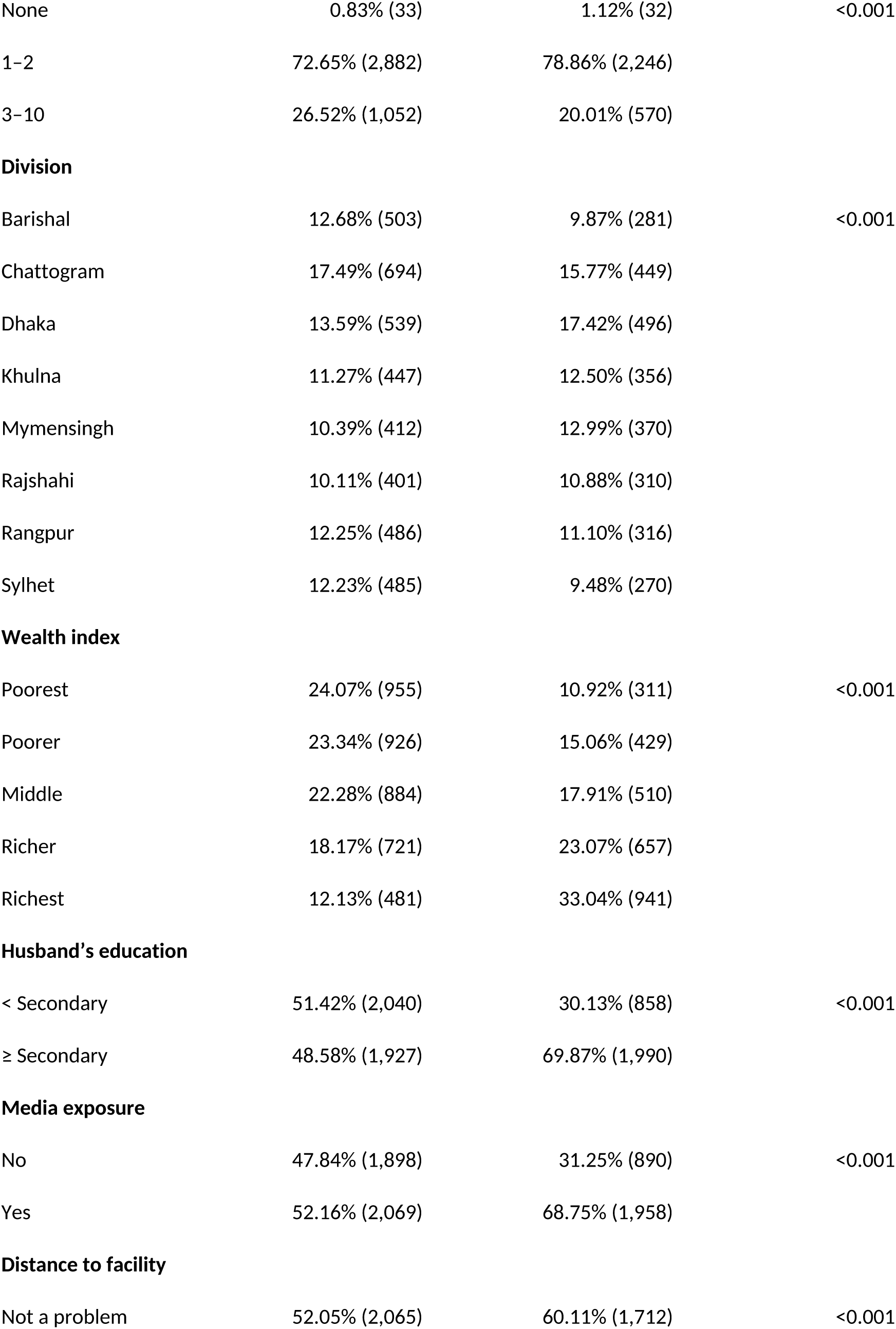
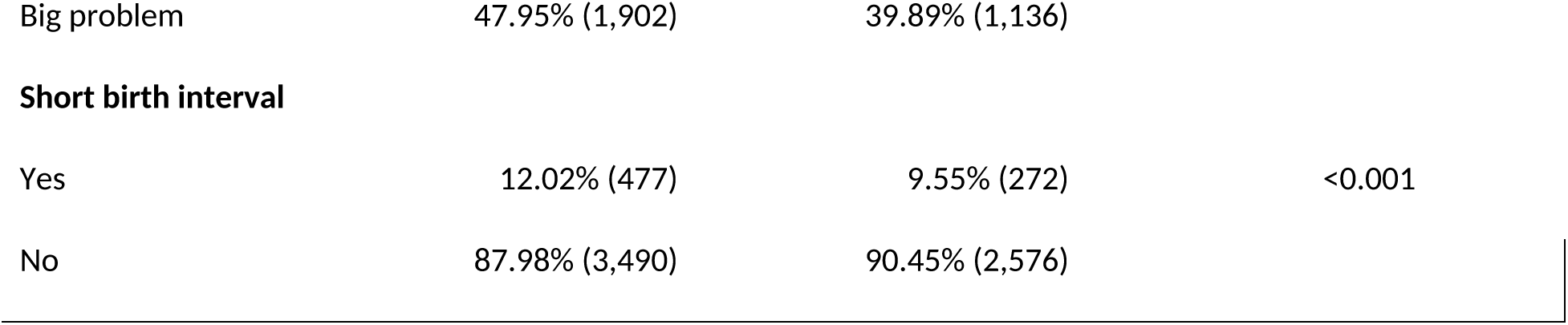
Association Between Maternal and Household Characteristics and Adequate Antenatal Care (ANC 4+)

| Variable | ANC < 4 (n = 3,967) | ANC ≥ 4 (n = 2,848) | p-value |
| --- | --- | --- | --- |
|  | % (n) | % (n) |  |
| Maternal education |  |  |  |
| < Secondary | 30.60% (1,214) | 16.64% (474) | <0.001 |
| ≥ Secondary | 69.40% (2,753) | 83.36% (2,374) |  |
| Age at first birth |  |  |  |
| ≤19 years | 66.85% (2,652) | 50.98% (1,452) | <0.001 |
| 20–40 years | 33.15% (1,315) | 49.02% (1,396) |  |
| Place of residence |  |  |  |
| Urban | 26.19% (1,039) | 44.03% (1,254) | <0.001 |
| Rural | 73.81% (2,928) | 55.97% (1,594) |  |
| Religion |  |  |  |
| Islam | 92.51% (3,670) | 89.43% (2,547) | <0.001 |
| Others | 7.49% (297) | 10.57% (301) |  |
| No. of living children |  |  |  |
| None | 0.83% (33) | 1.12% (32) | <0.001 |
| 1-2 | 72.65% (2,882) | 78.86% (2,246) |  |
| 3-10 | 26.52% (1,052) | 20.01% (570) |  |
| **Division** |  |  |  |
| Barishal | 12.68% (503) | 9.87% (281) | <0.001 |
| Chattogram | 17.49% (694) | 15.77% (449) |  |
| Dhaka | 13.59% (539) | 17.42% (496) |  |
| Khulna | 11.27% (447) | 12.50% (356) |  |
| Mymensingh | 10.39% (412) | 12.99% (370) |  |
| Rajshahi | 10.11% (401) | 10.88% (310) |  |
| Rangpur | 12.25% (486) | 11.10% (316) |  |
| Sylhet | 12.23% (485) | 9.48% (270) |  |
| **Wealth index** |  |  |  |
| Poorest | 24.07% (955) | 10.92% (311) | <0.001 |
| Poorer | 23.34% (926) | 15.06% (429) |  |
| Middle | 22.28% (884) | 17.91% (510) |  |
| Richer | 18.17% (721) | 23.07% (657) |  |
| Richest | 12.13% (481) | 33.04% (941) |  |
| **Husband's education** |  |  |  |
| < Secondary | 51.42% (2,040) | 30.13% (858) | <0.001 |
| ≥ Secondary | 48.58% (1,927) | 69.87% (1,990) |  |
| **Media exposure** |  |  |  |
| No | 47.84% (1,898) | 31.25% (890) | <0.001 |
| Yes | 52.16% (2,069) | 68.75% (1,958) |  |
| **Distance to facility** |  |  |  |
| Not a problem | 52.05% (2,065) | 60.11% (1,712) | <0.001 |
| Big problem | 47.95% (1,902) | 39.89% (1,136) |  |
| **Short birth interval** |  |  |  |
| Yes | 12.02% (477) | 9.55% (272) | <0.001 |
| No | 87.98% (3,490) | 90.45% (2,576) |  |

Women aged ≤19 years at first birth had better ANC coverage (51%) than those aged 20-40 years (49%) (p < 0.001). Rural women (56%) had higher ANC use than urban women (44%) (p < 0.001). ANC coverage increased with wealth, from 11% in the poorest group to 33% in the richest (p < 0.001) and was higher among those with media exposure (69%) than without (31%) (p < 0.001).

Women with 1–2 children had the highest ANC use (79%) compared to those with 3–10 children (20%) (p < 0.001). Regional variations existed, with Dhaka having the highest coverage (17%) and Sylhet the lowest (10%) (p < 0.001). ANC use was also higher among women who did not report distance as a big problem (p < 0.001) and among those without short birth intervals (p = 0.001).

### Model selection for PS estimation

Fig 1 presents the standardized mean differences (SMDs) for covariates before and after matching using propensity scores estimated by various machine learning models. Overall, covariate balance improved after matching across all models. Among them, the K-Nearest Neighbors (KNN) model demonstrated the best performance, with nearly all covariates achieving post-matching SMDs well below the conventional threshold of 0.1—indicating excellent balance. Logistic Regression also performed well, though a slightly higher number of covariates remained near the threshold. In contrast, the Support Vector Machine (SVM), Random Forest, and Neural Network models showed poorer performance in achieving balance; XGBoost offered moderate improvement.

**Fig 1.**
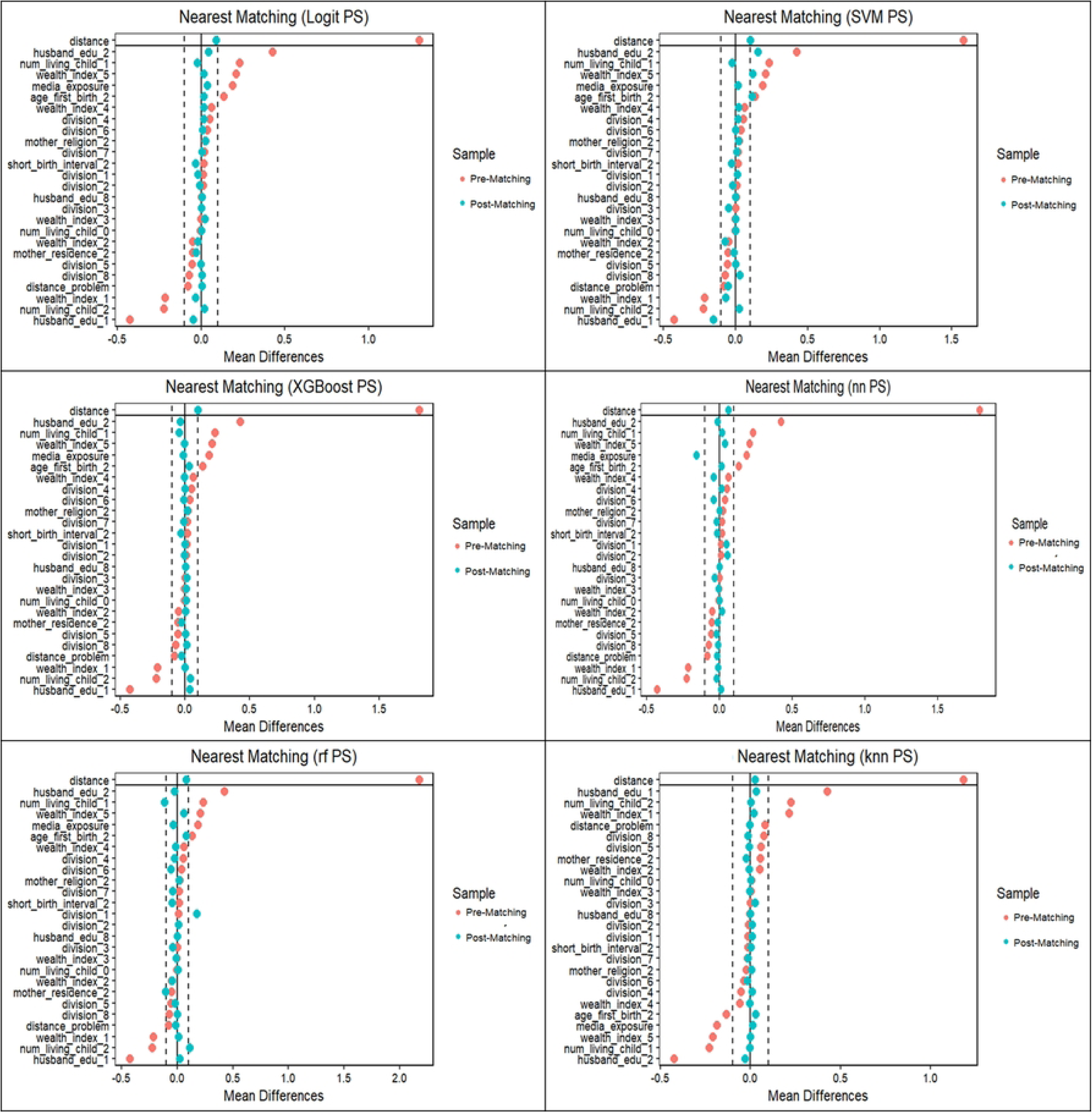
Covariate Balance Before and After Matching Using Propensity Scores Estimated by Different Machine Learning Models. Love plot displaying standardized mean differences for covariates before and after nearest-neighbor matching (caliper = 0.1), using propensity scores estimated via various machine learning models. The horizontal dashed line at 0.1 represents the commonly accepted threshold for acceptable covariate imbalance.

Fig 2 displays density plots of estimated propensity scores for treated and control groups, highlighting the degree of overlap (common support) before and after matching. While all models demonstrated acceptable overlap, Logistic Regression showed the highest overlap (0.93) and retained the largest number of matched pairs (1,480), followed by KNN, which achieved substantial overlap (0.89) and 1,132 matched pairs (Table 2).

**Fig 2.**
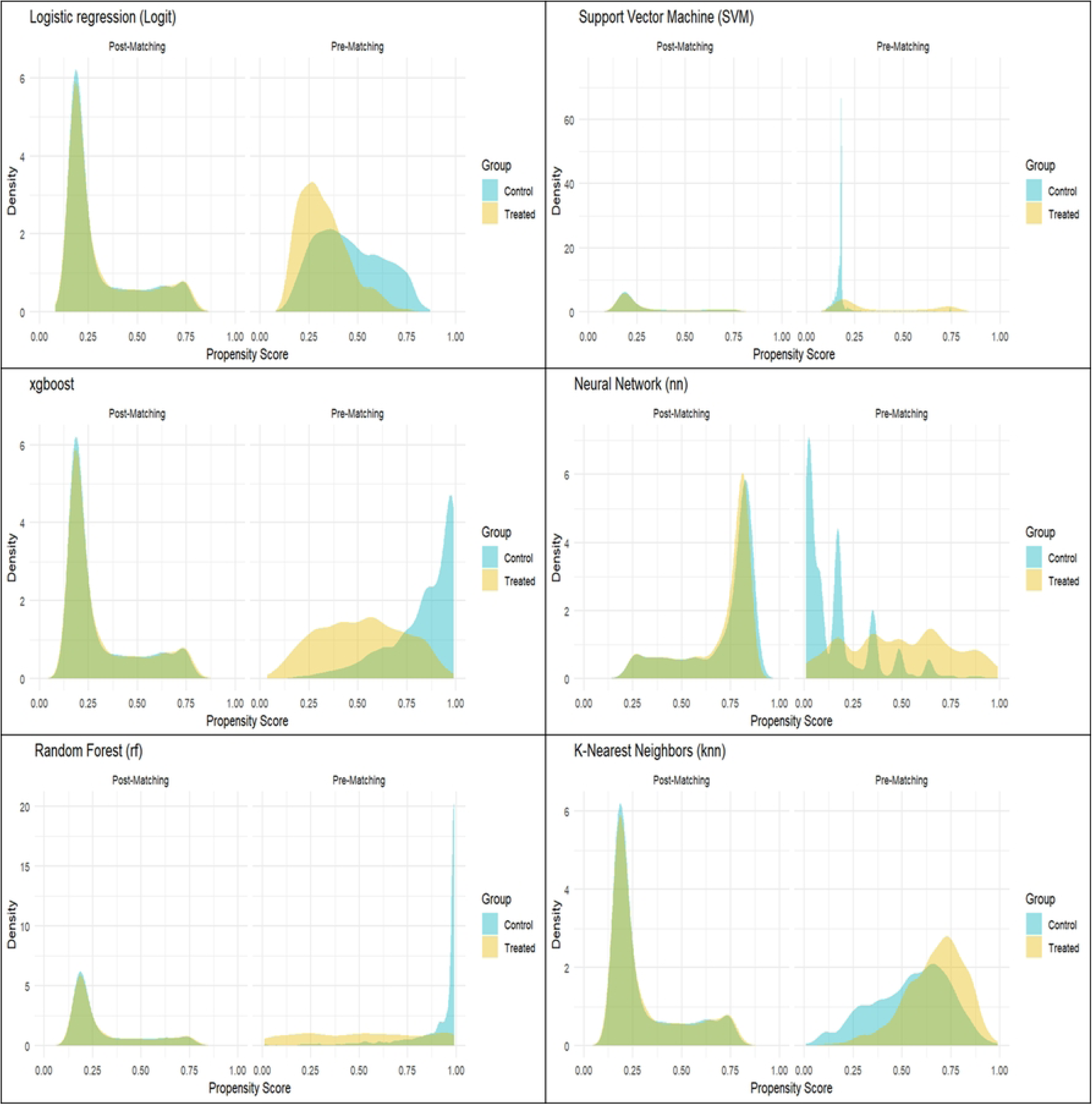
Propensity Score Overlap Between Treated and Control Groups Across Different Machine Learning Models. Density plots showing the distribution of estimated propensity scores for treated and control groups before and after matching, stratified by model. Overlap indicates the extent of common support achieved by each machine learning model, with higher overlap suggesting better suitability for causal comparisons.

**Table 2.** Comparison of Propensity Score Estimation Methods Based on Model Performance Metrics.

| Model | Matched Pairs | Overlap | Distance Balance | AUC |
| --- | --- | --- | --- | --- |
| Logit | 1480 | 0.93 | 0.09 | 0.80 |
| KNN | 1132 | 0.89 | 0.03 | 0.83 |
| SVM | 1287 | 0.89 | 0.10 | 0.82 |
| XGBoost | 1251 | 0.87 | 0.10 | 0.86 |
| Random Forest | 1132 | 0.78 | 0.08 | 0.86 |
| Neural Net | 1234 | 0.88 | 0.01 | 0.84 |

Despite Logistic Regression’s advantage in overlap and sample retention, KNN was selected as the preferred method for propensity score estimation in this study. This decision is based on its superior covariate balance and lower distance imbalance (0.03), which are critical when matching is used as a preprocessing step for non-parametric outcome modeling, such as causal forests (22). The non-parametric nature of KNN aligns well with causal forest assumptions and helps minimize residual confounding, offering a robust foundation for subsequent causal effect estimation.

### Treatment effect

The Average Treatment Effect (ATE) estimated using the full sample was 0.33 (95% CI: −0.11, 0.77) with a standard error of 0.22, yielding a non-significant p-value of 0.14 (Table 3). In comparison, the ATE estimated from the K-Nearest Neighbors (KNN) matched sample was smaller, at 0.006 (95% CI: −0.002 to 0.016), with a standard error of 0.005 and a p-value of 0.17, indicating no statistically significant treatment effect in either analysis.

**Table 3.** The average treatment effect (ATEs) and individual treatment effects (ITEs) from Causal Forest models.

| Type of estimates |  | Full sample | KNN matched |
| --- | --- | --- | --- |
| ATE | Average (95% CI) | 0.33 (-0.11, 0.77) | 0.006 (-0.002, 0.016) |
|  | Standard Error | 0.22 | 0.005 |
|  | P-values | 0.14 | 0.17 |
| ITE | Average | 0.078 | 0.006 |
|  | Median | 0.072 | 0.006 |
|  | Rang | -0.12-0.31 | 0.005-0.007 |

Regarding the Individual Treatment Effects (ITE), the average and median values were consistent within each sample but differed markedly between samples. The full sample demonstrated an average and median ITE of approximately 0.072, whereas the matched sample showed substantially lower values of 0.006 for both metrics. Additionally, the range of ITE values was narrower in the matched sample (0.005 to 0.007) compared to the full sample (−0.11 to 0.31), suggesting reduced variability following matching.

Fig 3 presents histograms of the estimated Individual Treatment Effects (ITEs) derived from the Causal Forest model using both the full sample and the matched (KNN propensity score) sample. Both distributions are approximately symmetric and centered around similar means. However, the ITE distribution from the matched sample exhibits a narrower spread, indicating reduced heterogeneity in treatment effects compared to the full sample.

**Fig 3.**
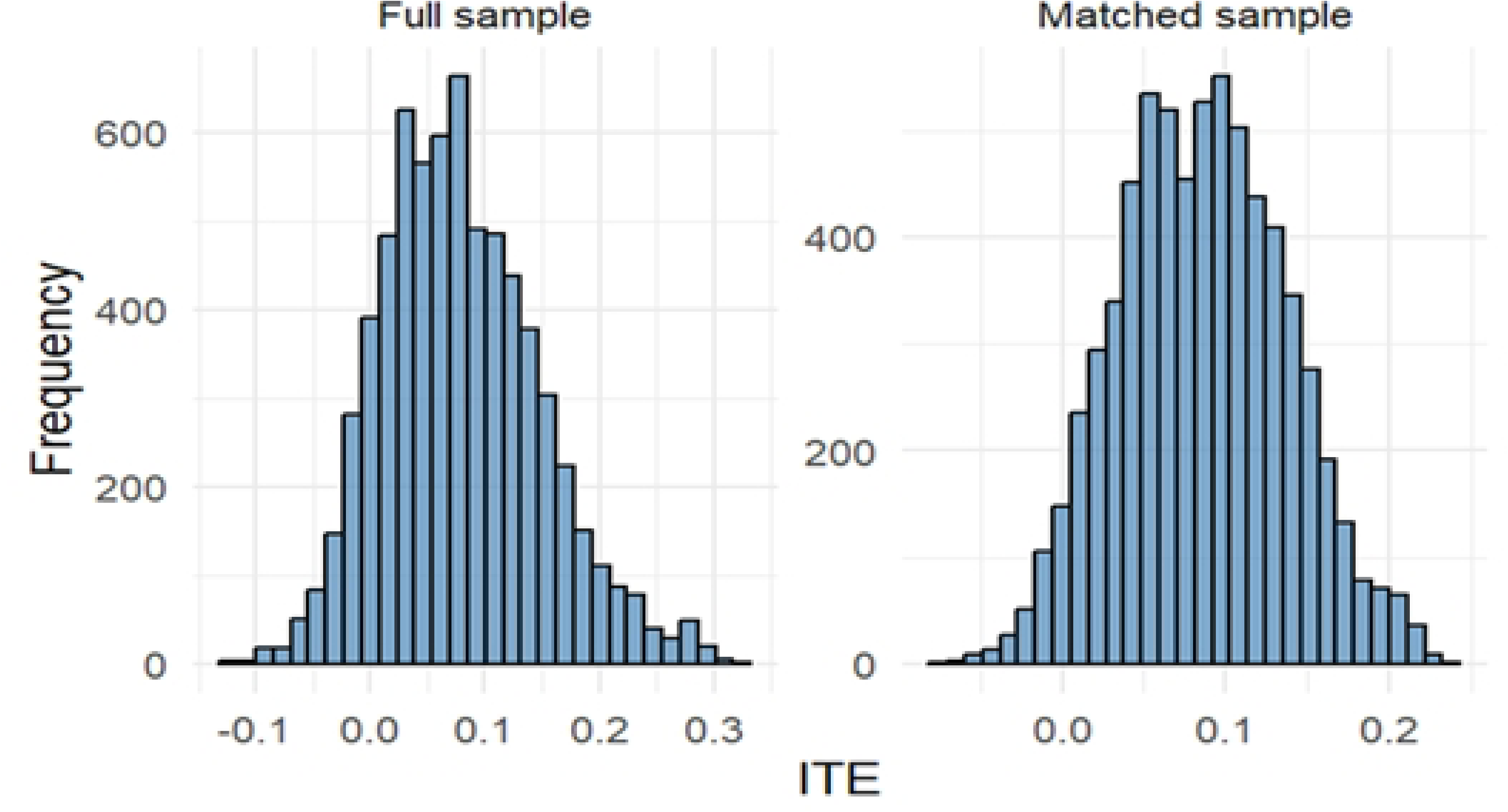
**Distribution of estimated Individual Treatment Effects (ITEs) from Causal Forest models using K-Nearest Neighbors (KNN) matching and non-matching**

### Heterogeneity analysis of Individual Treatment Effects (ITE)

Significant heterogeneity was observed in ITEs across maternal age at first birth, residence, husband’s education, media exposure, and wealth index. Specifically, women who gave birth at age 20–40 exhibited significantly higher average ITEs (mean = 0.09) compared to those who gave birth at or before age 19 (mean = 0.07), with a large effect size (*Cohen’s d* = –0.65, 95% CI: [–0.70, –0.60], *p* < 0.001) (Table 4, and S1. Supporting Information). Similarly, women residing in urban areas had higher ITEs (0.09) than those in rural settings (0.07), with a medium effect size (*d* = 0.43, 95% CI: [0.38, 0.48]).

**Table 4.** Heterogeneity in Average Individual Treatment Effects (ITEs) Across Subgroups.

| Variable | Average ITEs by Group | Statistical Test<br>(t/ANOVA); p-value | Effect Size<br>(Cohen's d /<br>Eta<sup>2</sup>) | 95%<br>Confidence<br>Interval (Effect<br>Size) |
| --- | --- | --- | --- | --- |
| Age at First Birth | ≤19 years: 0.0654;<br>20–40 years: 0.0936 | $t = -25.58; p < 0.001$ | $d = -0.65$ | [-0.70, -0.60] |
| Residence | Urban: 0.0893;<br>Rural: 0.0702 | $t = 16.49; p < 0.001$ | $d = 0.43$ | [0.38, 0.48] |
| Religion | Islam: 0.0762;<br>Other: 0.0814 | $t = -3.13; p = 0.0018$ | $d = -0.11$ | [-0.20, -0.03] |
| Husband's<br>Education | <Secondary: 0.0594;<br>≥Secondary: 0.0894 | $t = -27.28; p < 0.001$ | $d = -0.70$ | [-0.75, -0.65] |
| Media Exposure | No: 0.0689;<br>Yes: 0.0819 | $t = -11.83; p < 0.001$ | $d = -0.29$ | [-0.34, -0.24] |
| Short Birth Interval | No: 0.0809;<br>Yes: 0.0761 | $t = 3.37; p = 0.0008$ | $d = 0.11$ | [0.03, 0.18] |
| Wealth Index | Poorest: 0.0467;<br>Poorer: 0.0773;<br>Middle: 0.0834;<br>Richer: 0.0811;<br>Richest: 0.0916 | $F(4, 6810) = 209.9;$<br>$p < 0.001$ | $\eta^2 = 0.11$ | [0.10, 1.00] |
| No. of Living<br>Children | 0: 0.0685; 1–2: 0.0795; 3–<br>10: 0.0677 | $F(2, 6812) = 43.04;$<br>$p < 0.001$ | $\eta^2 = 0.01$ | [0.01, 1.00] |
| **Division** | Barishal: 0.1121;<br>Chattogram: 0.1009;<br>Dhaka: 0.0816;<br>Khulna: 0.0755;<br>Others: 0.0474–0.0674 | $F(7, 6807) = 265;$<br>$p < 0.001$ | $\eta^2 = 0.21$ | [0.20, 1.00] |

Cohen's d values: 0.01–0.19 =very small, 0.20–0.49=small, 0.50–0.79= medium, and 0.80+= large (25,26).
$\eta^2$ values: 0.01 = small, 0.06 = medium, 0.14+ = large (25).

A particularly strong gradient in ITEs was evident across levels of husband’s education, where women whose husbands had secondary or higher education had considerably higher ITEs (0.09 vs. 0.06; *d* = –0.70, 95% CI: [–0.75, –0.65], *p* < 0.001). Media exposure was also positively associated with ITEs (0.08 vs. 0.07; *d* = –0.29, *p* < 0.001).

While differences in ITEs by mother’s religion and short birth interval were statistically significant (*p* < 0.01), their effect sizes were small (*Cohen’s d* = –0.11 and 0.11, respectively), indicating limited practical significance.

The analysis of categorical variables using ANOVA showed substantial heterogeneity in ITEs across wealth quintiles (*η²* = 0.11), divisional regions (*η²* = 0.21), and number of living children (*η²* = 0.01). The largest variance in treatment effects was attributed to division, suggesting substantial geographical heterogeneity, while variation by number of living children was minimal.

### Sensitivity analysis

We performed a Rosenbaum bounds sensitivity analysis on the matched sample to assess the robustness of our estimated treatment effect to potential hidden bias from unmeasured confounders. The Wilcoxon signed-rank test p-value under no hidden bias (Γ= 1) was 0.0609, indicating a marginally non-significant treatment effect in the matched pairs analysis (Fig 4). Notably, the p-value remained constant at 0.0609 for Γ values ranging from 1 to 1.9, suggesting that the inference about the treatment effect is stable against moderate levels of unobserved confounding.

**Fig 4.**
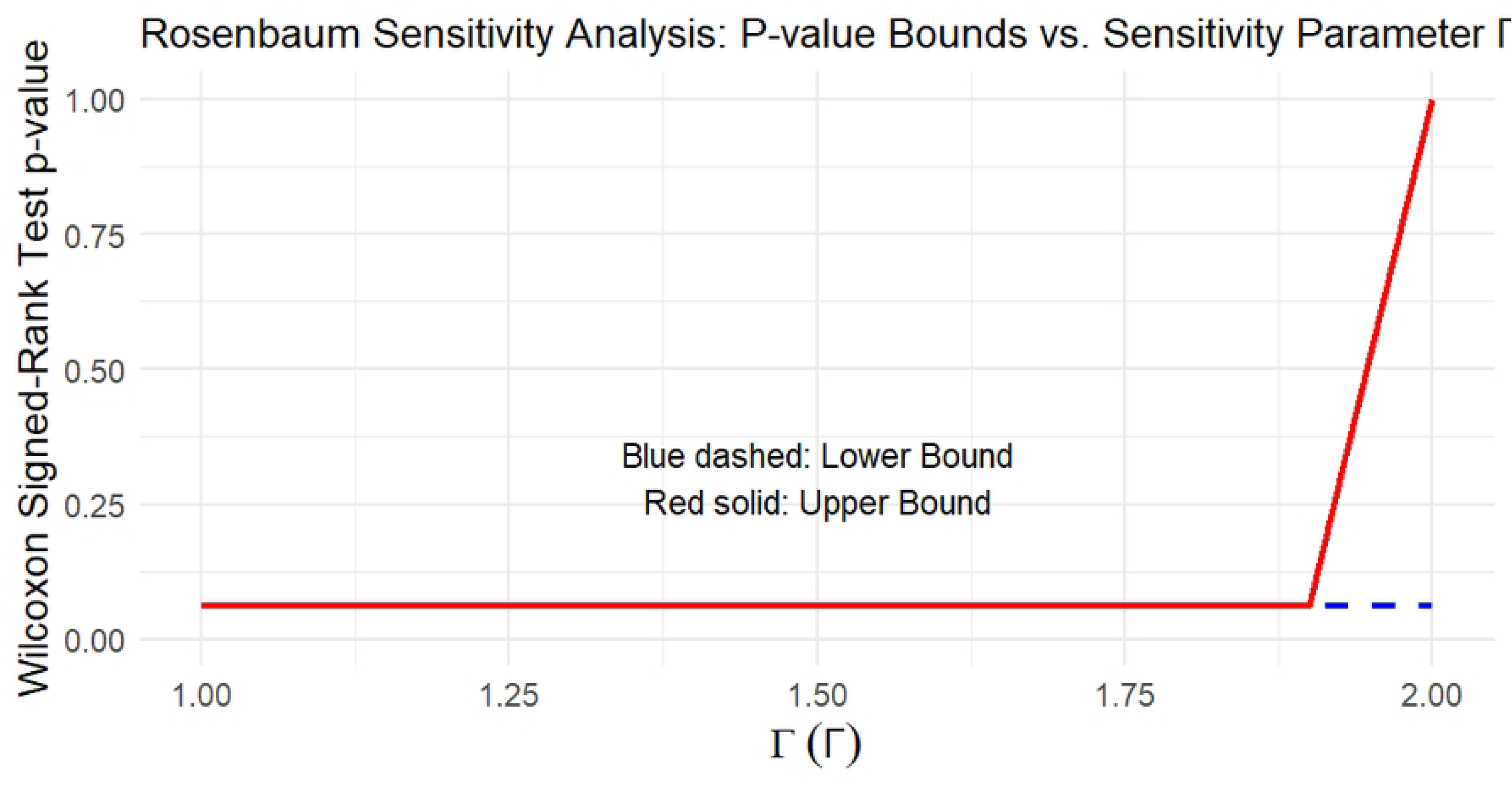
**Plot of Rosenbaum sensitivity analysis showing how the Wilcoxon signed-rank test p-values change as the sensitivity parameter Gamma (**𝜞**) increases**

At the highest tested level of hidden bias (Γ= 2), the bounds on the p-value ranged widely from 0 to 1, indicating that if the odds of differential assignment to treatment due to unobserved confounders doubled, the conclusion could shift from non-significant to significant.

## Discussion

In the unadjusted (unmatched) dataset, women with secondary or higher education showed significantly higher rates of adequate antenatal care (ANC) utilization at 83.4%, compared to only 16.6% among their less-educated counterparts. This stark contrast reflects longstanding national trends in Bangladesh, which also aligns with regional data from South Asia and Sub-Saharan Africa that consistently correlate maternal education to improved maternal and child health outcomes (2,3,5,27).

This substantial relationship is supported by multiple mechanisms. Education is believed to promote autonomy in reproductive health decision-making, improve cognitive ability to comprehend and act upon health information, and raise health literacy (28,29). Educated women are more likely to recognize the signs of pregnancy complications, attend ANC visits proactively, and adhere to provider recommendations. Moreover, education often correlates with greater media exposure, higher household income, and more equitable gender norms, all of which can facilitate health service utilization (30–33).

However, when adjusting for confounding through machine learning–based propensity score matching, the estimated Average Treatment Effect (ATE) using causal forest was minimal and not statistically significant (ATE = 0.006, 95% CI: −0.002 to 0.016, p = 0.17). This finding diverges from earlier studies employing traditional regression or simpler propensity score methods (5,34). For instance, Akter et al. (2024) found significant causal effects of maternal education on maternal health service uptake using various propensity score methods based on 1,300 recently delivered mothers in three sub-districts in Northern Bangladesh (1).

This contrast suggests that much of the observed association may be explained by confounding factors such as wealth, media exposure, place of residence, and partner’s education. The scenario we observed is that the maternal education is correlated with but not causally responsible for higher ANC use, once other socioeconomic and contextual variables are properly accounted for. Psaki et al. (2019) conducted a systematic review and meta-analysis of the causal effects of education on reproductive health outcomes and found small and inconsistent effects (32).

Crucially, a more complex picture is presented by our heterogeneity analysis employing Individual Treatment Effects (ITEs). Although maternal education seems to have a minor overall influence, its effects vary significantly within subgroups, which is consistent with recent research highlighting the value of disaggregated causation analysis in public health (35). For instance, women from wealthier households, those residing in urban areas, and those whose husbands had secondary or higher education consistently exhibited higher ITEs (See Table 4). This implies that maternal education exerts a synergistic effect when combined with enabling factors like economic security, spousal support, and geographic proximity to health facilities (2,5).

Other LMIC environments have reported similar conditional treatment effects. In Nepal, Furuta and Salway (2006) demonstrated that women’s education only increased service utilization when it was coupled with supportive social and familial structures, especially those that allowed for joint decision-making and spousal communication (36). Likewise, Ahmed et al. (2010) found in a cross-national study utilizing DHS data that maternal education had a greater impact on health-seeking behavior in settings where women had greater access to media, were financially independent, and had supportive spouses (37). In addition, the substantial effect sizes found in our subgroup analysis, particularly by wealth index (η² = 0.11), division (η² = 0.21), and husband’s education (Cohen’s d = 0.70), suggest that structural and contextual factors influence the benefits of education. Accordingly, education is not a “magic bullet” but a crucial part of a larger system of surroundings promoting health. The return on educational investments in terms of health outcomes may be low if these complementary factors are neglected (28,33,37).

The results of this study have important implications for maternal health policies in Bangladesh and other LMICs. Although encouraging female education remains a key component of public health strategy, mounting data indicate that education is insufficient to ensure appropriate ANC use and mother and child wellbeing. Complementary interventions are also required to address structural barriers, such as inadequate rural healthcare facilities, financial constraints, and power imbalances related to gender (2,38,39).

Improving healthcare access in rural areas is critical. The likelihood of receiving four or more ANC visits in Bangladesh decreases by 42% for every kilometer that separates one from a health facility, and care uptake is greatly increased by higher facility readiness (6). Programs for financial assistance, including conditional cash transfers (CCTs), have also shown promise; in LMICs, they raised ANC attendance by 5–45%, especially for women with lower incomes (40).

Male involvement in maternal health decision-making improves outcomes. Recent evidence from Bangladesh and Indonesia shows that partner support significantly enhances ANC attendance and shared health decisions, even among less-educated women (41). Further, given the geographical differences in treatment effects found in this study, targeted outreach, such as community education or mobile clinics, in underperforming areas like Sylhet and Barisal, may have a greater impact than uniform, national programs (42). Additionally, pairing education with media exposure and empowerment tools, including mobile health (mHealth) campaigns, has improved ANC use by enhancing awareness and autonomy among less-educated women (42).

## Strengths and limitations

The primary strength of our study is the incorporation of cutting-edge machine learning in propensity score estimation and the use of causal forests to explore treatment effect heterogeneity. Notwithstanding the study’s advantages, certain limitations should be noted. First, even with sophisticated causal inference techniques, the analysis depends on observational cross-sectional data, which leaves room for unobserved confounding. Second, the dataset lacked some potentially significant factors, including the distance (in kilometers) to medical facilities and the quality of care. Third, the outcome, ANC visits, was based on self-report, which may be prone to recall or social desirability bias.

## Conclusion

In conclusion, while maternal education is strongly associated with increased ANC utilization, its causal effect is small and varies across subpopulations. This suggests that the benefits of education may be contingent upon contextual factors that either constrain or enhance a woman’s ability to act on health knowledge. Integrating machine learning for matching and causal inference enables a more nuanced understanding of these dynamics, offering a pathway toward equity-informed and data-driven maternal health policy in low-resource settings like Bangladesh.

## Declarations

### Ethics approval and consent to participate

The Bangladesh Demographic and Health Survey (BDHS) obtained informed consent from all participants prior to data collection. Ethical clearance for conducting the BDHS was granted by the relevant authorities of the Government of Bangladesh. All procedures were carried out in accordance with national ethical guidelines and standards for research involving human participants.

### Availability of data and materials

The datasets used and/or analyzed during the current study are publicly available without restriction at https://dhsprogram.com/data/available-datasets.cfm. Additionally, the data and code files generated during the analysis are available from the corresponding author upon reasonable request.

## S1: Supporting Information

